# Inflammation and Oxidative Stress Markers in Type 2 Diabetes Patients with Advanced Carotid Atherosclerosis

**DOI:** 10.1101/2023.07.13.23292592

**Authors:** Louise Ménégaut, Aline Laubriet, Valentin Crespy, Damien Leuleu, Thomas Pilot, Kevin Van Dongen, Jean-Paul Pais de Barros, Thomas Gautier, Jean-Michel Petit, Charles Thomas, Maxime Nguyen, Eric Steinmetz, David Masson

**Affiliations:** Université Bourgogne, LNC UMR1231, Dijon, France; INSERM, UMR1231, Dijon, France; Université Bourgogne-Franche Comté, LipSTIC LabEx, Dijon, France; CHU Dijon, Laboratory of Clinical Chemistry, Dijon, France; CHU Dijon, Department of Cardiovascular and Thoracic Surgery, Dijon, France; CHU Dijon, Department of Endocrinology and metabolic diseases, Dijon, France; CHU Dijon Department of Anesthesiology and Intensive Care, Dijon, France; Lipidomic Analytic Platform, Université Bourgogne Franche-Comté, Dijon, France

## Abstract

**Background:** Type 2 diabetes mellitus (T2DM) is a major global health issue and a significant risk factor for atherosclerosis. Atherosclerosis in T2DM patients has been associated with inflammation, insulin resistance, hyperglycemia, dyslipidemia, and oxidative stress. Identifying molecular features of atherosclerotic plaques in T2DM patients could provide valuable insights into the pathogenesis of the disease

**Aim:** The MASCADI (Arachidonic Acid Metabolism in Carotid Stenosis Plaque in Diabetic Patients) study aimed to investigate the increase of 2-arachidonoyl-lysophatidylcholine (2-AA– LPC) in carotid plaques from T2DM and control patients and to explore its association with plaque vulnerability as well as with blood and intra-plaque biomarkers altered during diabetes.

**Results:** In a population of elderly, polymedicated patients with advanced stage of atherosclerosis, we found that T2DM patients had higher systemic inflammation markers, such as high-sensitivity C-reactive protein (hsCRP) and IL-1β, higher levels of oxysterols, increased triglyceride levels, and decreased HDL levels as compared to control patients. Furthermore, 2-AA-LPC was significantly enriched in plaques from diabetic patients, suggesting its potential role in diabetic atherosclerosis. Interestingly, 2-AA-LPC was not associated with systemic markers related to diabetes, such as hsCRP, triglycerides, or HDL cholesterol. However, it was significantly correlated with the levels of inflammatory markers within the plaques such as lysophospholipids and 25-hydroxycholesterol, strengthening the link between local inflammation, arachidonic acid metabolism and diabetes.

**Conclusion:** Our study is in line with a key role for inflammation in the pathogenesis of diabetic atherosclerosis and highlights the involvement of 2-AA-LPC. Further research is needed to better understand the local processes involved in the alteration of plaque composition in T2DM and to identify potential therapeutic targets.

## Introduction

Type 2 diabetes mellitus (T2DM) is a growing global health concern, affecting millions of people worldwide. It has been widely recognized as an independent risk factor for atherosclerosis, a major contributor to cardiovascular diseases (CVDs) which represent the leading cause of morbidity and mortality in patients with T2DM.^1^ Despite extensive research on this field, the underlying mechanisms involved in the development and progression of atherosclerosis in patients with T2DM remain elusive due to their multiplicity and intricate relationships. Indeed, many factors have been implicated in the pathogenesis of atherosclerosis in T2DM, including inflammation, insulin resistance, hyperglycemia, dyslipidemia, and oxidative stress. ^2–4^ Hyperglycemia, for instance, has been associated with increased production of advanced glycation end-products (AGEs), which can contribute to endothelial dysfunction and plaque formation.^5, 6^. Additionally, diabetic dyslipidemia, characterized by elevated triglycerides and low-density lipoprotein (LDL) cholesterol levels, as well as reduced high-density lipoprotein (HDL) cholesterol levels, has been linked to increase plaque vulnerability and accelerated atherosclerosis in T2DM patients^7^. Oxidative stress, caused by an imbalance between the production of reactive oxygen species (ROS) and the antioxidant defense mechanisms, has also been implicated in the development of atherosclerosis in T2DM patients.^8^ Elevated ROS levels are well known to promote inflammation, endothelial dysfunction, and plaque instability, thereby exacerbating the atherosclerotic process.^9^

In parallel to these advances in understanding the factors contributing to atherosclerosis in T2DM, several studies indicate that atheroma plaques in diabetic patients possess unique characteristics in terms of morphology, vulnerability, composition, and calcification ^10–13^. Therefore, identifying molecular features of atherosclerotic plaques in T2DM patients could provide valuable insights into the pathogenesis of the disease and lead to the development of more targeted therapeutic strategies. Recent lipidomic analyses of atheroma plaques have revealed specific signatures associated with diabetes, such as increased free cholesterol content and altered levels of specific phospholipids and lysophospholipids (LPLs). ^14–16^ Again, the mechanisms behind these alterations and their implications for atherosclerosis in T2DM patients remain unclear. In a recent cohort study of 79 patients, we observed a specific enrichment of 2-arachidonoyl-lysophatidylcholine (2-AA-LPC) among other LPC in the plaques of diabetic patients, suggesting an important role for this molecule and more generally for arachidonic acid metabolism in atherosclerosis associated with diabetes.^17^ However, in a subsequent study, we did not observe any differences in AA derived mediators such as leukotriene B4 (LTB4), thromboxane B2 (TxB2) or prostaglandin E2 (PGE2) in carotid plaques from control or T2DM patients.^18^ The aim of the MASCADI (Arachidonic Acid Metabolism in Carotid Stenosis Plaque in Diabetic Patients) study was not only to confirm the increase in 2– AA-LPC in a larger cohort (101 T2DM and 99 control patients) but also to explore the association of this molecule with plaque vulnerability as well as with systemic and local biomarkers that are altered during the course of diabetes. Notably, we examined the relationships between 2-AA-LPC levels and inflammatory markers, triglycerides, glycated hemoglobin (HbA1c), and oxidative stress markers such as oxysterols.

### Material and methods

The MASCADI protocol was reviewed and approved by the regional Ethics Committee (Comité de Protection des Personnes Est, Dijon, France CPP Est III, CHRU Nancy, N° 2017-A00022-51) and recorded on ClinicalTrials.gov (clinical registration number: NCT03202823). Two-hundred– and-one patients were enrolled. As described previously, patients enrolled in the present study were admitted to the Department of Cardiovascular Surgery at the University Hospital of Dijon (Burgundy, France) for the surgical treatment of an atheromatous internal carotid artery (ICA) stenosis whether symptomatic or not.^18^ A stenosis was considered symptomatic if stroke or transient ischemic attack (TIA) attributed to the ICA lesion occurred within the previous 6 months before intervention. According to the trial category (Interventional research that does not involve drugs or non-CE-marked medical devices and that involves only minimal risks and constraints for the patient) and the ethics committee, all patients received written information note on the study. Oral consent was obtained from the patient and an attestation of the patient’s oral consent was signed by the investigator physician and countersigned by the patient, according to the french law in this type of trial. Blood serum samples and EDTA total blood samples were collected in parallel on the day of surgery. For all patients, carotid endarterectomy (CEA) was performed within the carotid bulb, with en-bloc removal of the entire plaque, and resulting samples were frozen at –80 °C prior to lipidomic analysis.

### Clinical data, imaging plaque characteristics, and biological analysis

Demographics, vascular risk factors (hypertension, active smoking, type 2 diabetes, dyslipidemia with detailed treatment), chronic renal insufficiency and symptomatic stenosis were collected. Patients were divided into groups according to their diabetic status: control group and diabetic group. Preoperative plaque imaging assessment included for all patients doppler ultrasound evaluating plaque echolucency and degree of stenosis according to NASCET measures, and CT angiographies (CTA) describing the presence of plaque ulceration and calcification. Glycated hemoglobin A1c (HbA1c) was measured by high pressure liquid chromatography on a Tosoh G8 analyzer (Tosoh Bioscience, Tokyo, Japan), C reactive protein (CRP), troponin I, creatinine, total cholesterol, HDL cholesterol and triglycerides were determined on a Dimension Vista analyzer using dedicated reagents (Siemens). Serum IL-1β levels were determined using a high-sensitivity enzyme-linked immunosorbent assay (ThermoFisher Scientific) with a limit of quantification (LOQ) of 0.05 pg/mL

### Lysophospholipid quantitation by LC-MS2

For lipidomic analysis, carotid plaques were opened longitudinally and core samples of atheroma plaques were harvested by using 3 mm biopsy punches (Kai Medical, Solingen, Germany). Sites of harvesting were normalized between the samples by selecting the plaque area corresponding to the maximal stenosis. Plaque biopsies (2–10 mg) were crushed in an Omni Bead Ruptor 24 apparatus (Omni International, Kennesaw, USA) with 1000 µL of phosphate buffer saline and circa twenty 1.4 mm OD zirconium oxide beads (S=6.95 m/s, T=30s, C = 3; D = 10s). Lipids corresponding to 1 mg of tissue were spiked with a mix containing 1 mg of di14:0-PC, 1 mg of di14:0-PE, 1 mg of d18:1/17:0– SM and 0.1 mg of 14:0-LPC used as internal standards. Phospholipids and lysophospholipids were analyzed by LC-MS/MS using a 6460 mass spectrometer (Agilent Technologies) equipped with an electrospray ionization source and MassHunter data system. The liquid chromatography was performed on a ZorBAX Eclipse SB-C18 (2.1 mm x 50 mm, 1.8 mm) (Agilent Technologies, Santa Clara, CA, USA).

### Plasma and plaque oxysterol quantitation by gas chromatography mass spectrometry (GC– MS)

Homogenates from plaques were crushed with NaCl 0.9% to obtain a final concentration of 0.33 mg tissue per ml. A volume of sample corresponding to 15 mg of tissue was saponified for 45 min at 56°C with 60 μl of potassium hydroxide 10 mol·L^−1^ and 1.2 ml of ethanol-BHT (50 mg·L^−1^) containing 20 μl of standard-intern mix (μg per sample: 0.2 (25-OH-cholesterol-d6); 0.4 (27-OH-cholesterol-d6); 0.5 (7α-OH-cholesterol-d7); 0.5 (7β-OH-cholesterol-d7); 1 (7-keto– cholesterol-d7)). Sterols were extracted with 5 ml of hexane and 1 ml of water. After evaporation of the organic phase, sterols were derivatized with 100 μl of a mixture of bis (trimethylsilyl)trifluoroacetamide/trimethylchlorosilane 4/1 v/v for 1 h at 80°C. After evaporation of the silylating reagent, 100 μl of hexane were added. Analysis of trimethylsilyl ethers of sterols was performed by GC–MS in a 7890A gas chromatograph coupled with a 5975C Mass Detector (Agilent Technologies). Separation was achieved on a HP-5MS 30 m × 250 μm column (Agilent Technologies, Santa-Clara, USA) using helium as carrier gas and the following GC conditions: injection at 250°C using the pulsed split, oven temperature programme: initial temperature 150°C up to 280°C at a rate of 15°C·min^−1^, up to 290°C at a rate of 1°C·min^−1^ for 2 min. The MSD was set up as follow: EI at 70 eV mode, source temperature at 230°C. Data were acquired in SIM mode using following quantitation ions (m/z): 131.1 for 25-OH-cholesterol; 137.1 for 25-OH-cholesterol d6; 417.4 for 27-OH– cholesterol; 423.4 for 27-OH cholestrol-d6; 456.4 for 7α–7β cholesterol; 463.5 for 7α–7β cholesterol-d7; 472.4 for 7-keto-cholesterol; 479.4 for 7-keto-cholesterol d7.

### Statistical analysis

We compared patients with diabetes to patient without diabetes. Normality was assessed using the Shapiro–Wilk test. Accordingly, quantitative data were presented as medians [interquartile range] or means (standard deviation) if normally distributed. Groups were compared with a Student’s t-test or the Kruskall–Wallis nonparametric test. When indicated, significance was corrected for multiple comparison by the Benjamini-Hochberg procedure. Qualitative data are presented as absolute value and percentage and compared by chi-squared or Fisher’s exact tests, as appropriate. Correlations were evaluated with the Spearman correlation method. Missing data were considered to be at random and were omitted in the analysis. R studio was used for statistical analysis.

## Results

### Patient characteristics and biological parameters in type 2 diabetic and control patients

The baseline characteristics are presented in Table 1. T2DM and control patients displayed similar demographic characteristics. Diabetic patients had a higher prevalence of hypertension and a higher BMI. No differences were found regarding the frequency of symptomatic plaques or major plaque characteristics (echogenicity, ulceration). However, the frequency of plaques with high levels of calcifications was higher in T2DM patients. The degree of stenosis was also significantly increased in diabetic patients. Regarding the biological parameters, T2DM patients presented relatively balanced glycemic parameters (Hba1c: 5.74% Vs 6.79% p<0.0001). As expected, there was a significant increase in plasma triglyceride levels associated with a decrease in HDL levels in diabetic patients. There were minor differences in total cholesterol and LDL cholesterol levels, but as expected the majority of the patients in both groups were under statin treatment.

**Table 1:** Baseline characteristics. Data are expressed as mean +/– S.D. (for normal distribution), medians [25^th^-75^th^ percentiles] or % of the population. Groups were compared with a student’s test or Kruskall-Wallis non-parametric test; qualitative data were compared using chi-squared or Fischer’s test.

|  | Controls<br>(*n*=98) | Diabetics<br>(*n*=101) | p-value |
| --- | --- | --- | --- |
| Age (years) | 74.0 [62.2;79.0] | 72.0 [65.0;78.0] | 0.766 |
| Gender (M/F) | 72/26 | 77/24 | 0.653 |
| BMI (kg/m<sup>2</sup>) | 26.0 [23.0;28.0] | 27.4 [25.0;30.0] | 0.001 |
| Active Smoking | 28 (28.6%) | 24 (23.8%) | 0.440 |
| HTA | 80 (81.6%) | 95 (94.1%) | 0.007 |
| Renal insufficiency | 11 (11.2%) | 12 (11.9%) | 0.885 |
| Diabetic treatment |  |  |  |
| No treatment or MHD |  | 9 (10%) | - |
| Biguanid |  | 41 (45%) | - |
| Sulfonylurea |  | 12 (13%) | - |
| Insulin |  | 16 (17%) | - |
| DDP-4 inhibitor |  | 5 (5.4%) | - |
| other |  | 4 (4.4 %) | - |
| Hypolipidemic treatment |  |  |  |
| Statin | 53 (57%) | 58 (63%) |  |
| Symptomatic lesions | 38 (38.8%) | 42 (41.6%) | 0.686 |
| Hyperechoic | 29 (29.6 %) | 36 (35.6 %) | 0.632 |
| Hypoechoic | 18 (18.4 %) | 15 (14.9 %) | 0.505 |
| NASCET | 75.0 [65.0;85.0] | 80.0[70.0;87.0] | 0.005 |
| Plaque Ulceration | 13 (13.3%) | 15 (14.9%) | 0.748 |
| Calcifications |  |  | 0.104 |
| 1 | 14 (14.3%) | 17 (16.8%) |  |
| 2 | 75 (76.5%) | 65 (64.4%) |  |
| 3 | 9 (9.2%) | 19 (18.8%) |  |
| HbA1c | 5.74 [5.48;5.97] | 6.79 [6.34;7.48] | <0.001 |
| Creatinine.μmol/L | 74.9 [60.9;89.0] | 71.8 [57.4;85.4] | 0.305 |
| Total cholesterol mmol/L | 3.88 [3.24;4.44] | 3.56 [2.83;4.21] | 0.043 |
| Triglycerides mmol/L | 1.36 [0.94;1.82] | 1.67 [1.14;2.36] | 0.017 |
| LDLc mmol/L | 2.04 [1.49;2.66] | 1.98 [1.46;2.54] | 0.407 |
| HDLc mmol/L | 1.13 (0.34) | 0.96 (0.28) | <0.001 |

### Systemic inflammation and oxidative stress markers in the study population

In order to explore systemic inflammation and oxidative stress markers, levels of high-sensitivity C-reactive protein (hs-CRP), IL-1β, and serum concentrations of oxysterols were measured in our population. The 7-oxidized forms are primarily formed during nonenzymatic oxidative processes and are markers of oxidative stress in diabetes^19, 20^, whereas 27-hydroxycholesterol and 25-hydroxycholesterol are produced enzymatically, with 25-hydroxycholesterol being actively produced upon inflammatory response. Interestingly, although the population was mainly beyond 70 years old and affected by advanced stage atherosclerosis, a significant difference was found regarding inflammatory markers, with increased levels of hs-CRP and IL-1β in type 2 diabetic patients (Table 2, Figure 1A-B). 7β-hydroxycholesterol and 7α-hydroxycholesterol levels were significantly increased in T2DM while there was no difference for other oxysterols (Table 2). Interestingly, hsCRP levels were positively associated with plasma triglyceride levels, both in the general population and among diabetic patients (r=0.2746, p=0.0057 in T2DM and r=0.348, p=0.0004 in controls) (Figure 1C).

**Figure 1:**
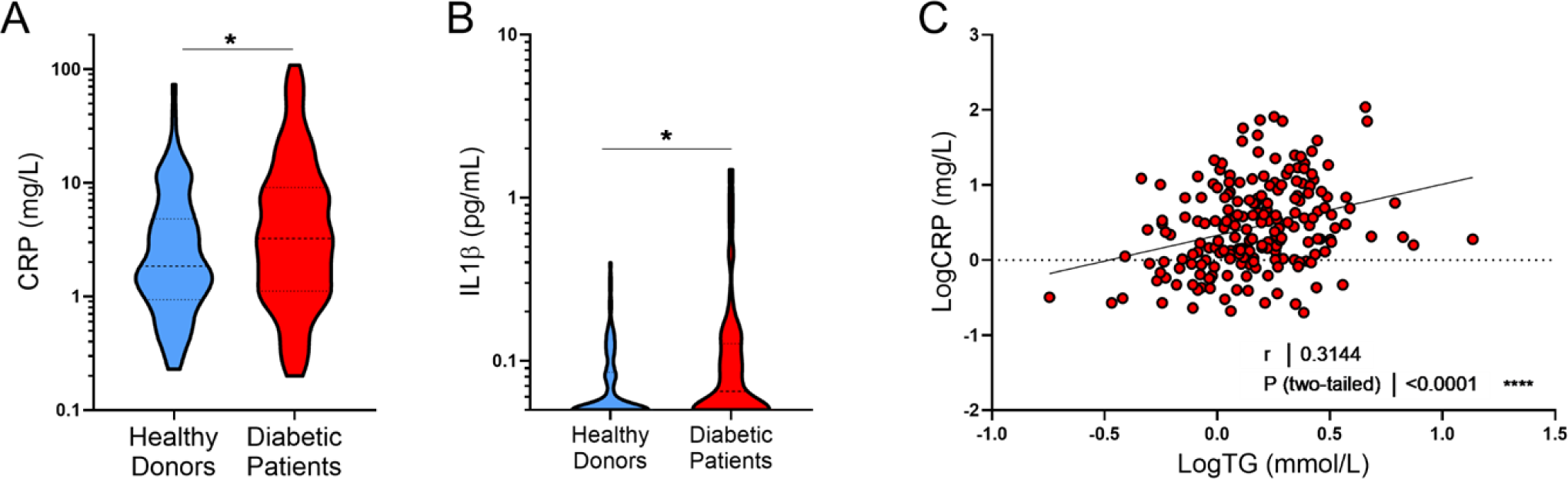
A Violin plot of C-reactive protein (CRP) levels in control and T2DM patients **B:** Violin plot of IL-1β levels in control and T2DM patients. **C**: Correlation between plasma HsCRP levels and triglycerides (R and P values were obtained by the Spearman rank test).

**Table 2:** Plasma oxysterols levels and inflammation biomarkers in controls and diabetic patients. Data are expressed as medians [25th-75th percentiles]. Groups were compared with a Mann-Whitney non-parametric test;

|  | Controls | Type 2 diabetics | p.overall |
| --- | --- | --- | --- |
| Oxysterols (mmol/mol cholesterol) |  |  |  |
| 7α-Hydroxycholesterol | 0.31 [0.21;0.42] | 0.36 [0.26;0.46] | 0.029 |
| 7β-Hydroxycholesterol | 0.39 [0.26;0.52] | 0.47 [0.33;0.66] | 0.040 |
| 7-ketocholesterol | 0.85 [0.59;1.37] | 0.90 [0.70;1.38] | 0.166 |
| 25-hydroxycholesterol | 0.14 [0.11;0.19] | 0.14 [0.10;0.22] | 0.854 |
| 27-hydroxycholesterol | 0.04 [0.03;0.05] | 0.04 [0.03;0.06] | 0.652 |
| Hs CRP (mg/L). | 1.85 [0.96;4.75] | 3.24 [1.17;8.75] | 0.028 |
| IL-1β (ng/l) | 0.05 [0.05;0.08] | 0.06 [0.05;0.12] | 0.017 |

### Enrichment of 2-AA-LPC in carotid plaques of diabetic patients

We had previously found an increase in the relative proportion of 2-AA-LPC within other lysophosphatidylcholines in carotid plaques of type 2 diabetic patients.^17^ In the present study, we could confirm these alterations in a larger cohort of patients. LPC levels were measured as previously described. As shown in table 3 while total LPC levels were not significantly different in the two populations, we confirmed the relative increase of 2-AA-LPC in diabetic patients (0.37% in control Vs 0.45% in T2DM patients p= 0.013), thus confirming the interest of this molecule in the context of diabetic atheroma. As with plasma, we also measured oxysterols in the plaques, focusing on the most abundant oxysterols (25-hydroxycholesterol, 27-hydroxycholesterol, 7α-hydroxycholesterol, 7β-hydroxycholesterol, and 7-ketocholesterol). Levels of oxysterols within the plaque have also been associated with inflammation, oxidative stress and plaque evolution ^21–24^. As indicated in Table 3, a trend toward an increase in 7-β hydroxycholesterol was found in diabetic patients, while other species were not significantly altered.

**Table 3:** Lysophosphatidylcholine and oxysterols in carotid plaques from control and type 2 diabetic patients. Data are expressed as mean +/– S.D. (for normal distribution), medians [25th-75th percentiles]. Groups were compared with a Student’s test or Kruskall-Wallis non-parametric test;

|  | Controls<br>*N=98* | Type 2 Diabetics<br>*N=101* | p.overall |
| --- | --- | --- | --- |
| Total LPC<br>(mmol/mol cholesterol) | 5.23 [3.05;6.93] | 4.84 [3.36;8.10] | 0.941 |
| LPC relative levels |  |  |  |
| 16.0.LPC.sn1 | 40.5 (2.81) | 40.8 (3.10) | 0.543 |
| 16.0.LPC.sn2 | 6.84 [6.07;7.46] | 6.82 [6.12;7.55] | 0.977 |
| 18.2.LPC.sn1 | 5.38 [4.08;6.84] | 5.26 [3.89;6.89] | 0.829 |
| 18.2.LPC.sn2 | 1.06 [0.74;1.37] | 1.02 [0.80;1.44] | 0.946 |
| 18.1.LPC.sn1 | 10.0 (2.04) | 9.83 (1.81) | 0.505 |
| 18.1.LPC.sn2 | 1.83 [1.54;2.23] | 1.88 [1.52;2.12] | 0.870 |
| 18.0.LPC.sn1 | 26.2 (4.18) | 25.5 (4.28) | 0.294 |
| 18.0.LPC.sn2 | 4.52 [3.92;5.44] | 4.49 [3.65;5.29] | 0.333 |
| 20.4.LPC.sn1 | 1.50 [1.13;1.98] | 1.55 [1.19;2.26] | 0.132 |
| 20.4.LPC.sn2 | 0.37 [0.26;0.62] | 0.45 [0.34;0.75] | 0.013 |
| 20.3.LPC.sn1 | 0.84 [0.65;1.07] | 0.87 [0.68;1.21] | 0.201 |
| 20.3.LPC.sn2 | 0.20 [0.14;0.26] | 0.19 [0.15;0.30] | 0.337 |
| Oxysterols (mmol/mol cholesterol) |  |  |  |
| 27-Hydroxycholesterol | 3.33 [2.02;7.25] | 3.35 [1.84;6.74] | 0.945 |
| 25-Hydroxycholesterol | 0.39 [0.28;0.62] | 0.43 [0.30;0.75] | 0.273 |
| 7α-Hydroxycholesterol | 0.75 [0.44;1.11] | 0.78 [0.49;1.47] | 0.246 |
| 7β-Hydroxycholesterol | 0.94 [0.64;1.27] | 1.11 [0.70;1.76] | 0.063 |
| 7-ketocholesterol | 2.03 [1.16;3.88] | 1.93 [1.11;3.94] | 0.940 |

### Independent association of 2-AA-LPC with T2D risk

After confirming the increase of 2-AA-LPC within the plaques of diabetic patients, we wanted to ensure that these molecules were independently associated with the risk of type 2 diabetes through a multivariate logistic regression, including the altered lipid markers within the plaques (7β-hydroxycholesterol, 2-AA-LPC) and the plasma markers modified during diabetes (HsCRP, triglycerides, HbA1c, and 7β-hydroxycholesterol). In all cases, 2-AA-LPC was positively and independently associated with the risk of diabetes, whether HbA1c was incorporated or not in the model (Figure 2 AB).

**Figure 2:**
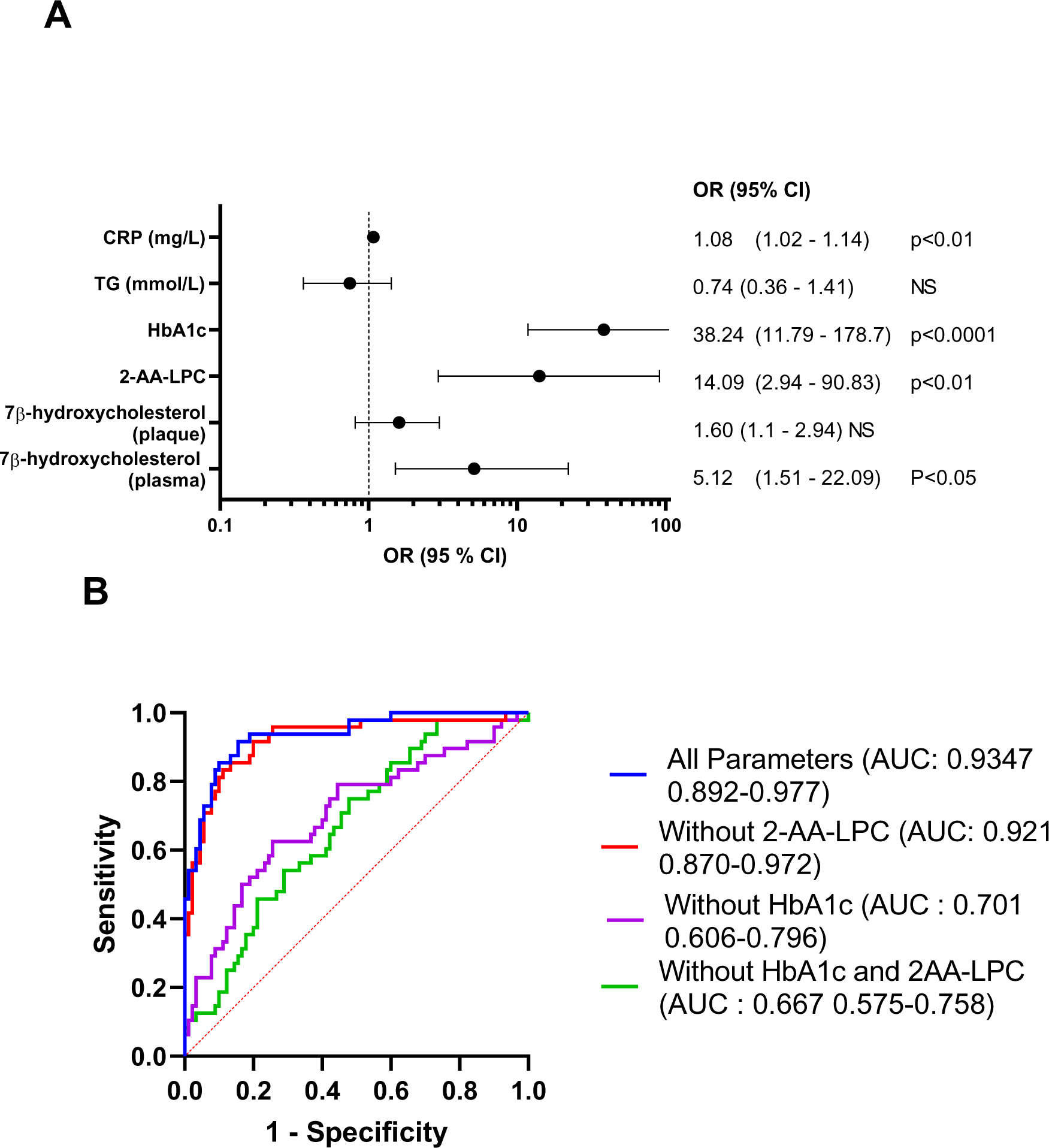
A: Forest plots showing odds ratio values and 95% confidence intervals for multivariable logistic regression analysis of diabetes and biological variables. **B:** Receiver operating characteristic curve (ROC) for the prediction of type 2 diabetes based on the selected biological parameters. (AUC: Area under the Curve)

### Exploring the Mechanisms behind Increased 2-AA-LPC in T2D Plaques

Finally, in order to understand the mechanisms associated with the increase of 2-AA-LPC in the plaques of diabetic patients, we examined the association, through a correlation matrix, between 2AA-LPC, the biomarkers measured within the plaque (LPC and oxysterols) with plasma markers related to hyperglycemia (HbA1c), dyslipidemia (triglycerides), inflammation (CRP), and oxidative stress (7-β-OH-cholesterol). Interestingly, no correlation was found between the plasma markers and the markers measured within the plaque, suggesting that the alterations in the plaque composition are mainly driven by local metabolic and inflammatory processes (figure 3A). As for the plaque markers, 2-AA-LPC appeared to be primarily associated with markers related to inflammation (total levels of LPC, 27-hydroxycholesterol, 25-hydroxycholesterol) and negatively with oxidative stress markers (Figure 3A). Interestingly, a strong correlation was found between the levels of 25– hydroxycholesterol and the levels of 2-AA-LPC, suggesting related production pathways (figure 3 AB).

**Figure 3:**
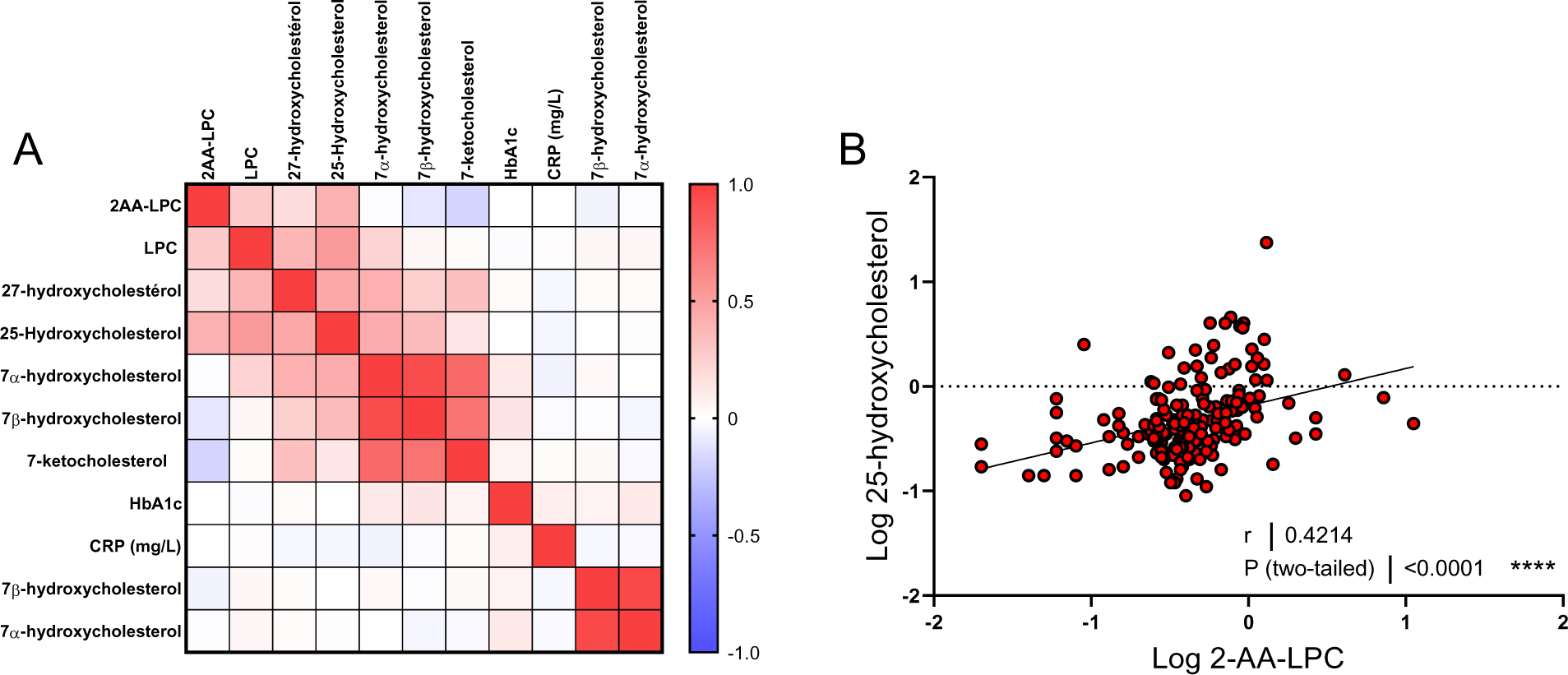
A: Correlation matrix of 2-AA-LPC with systemic and carotid plaque biomarkers. Correlations were determined by Spearman’s rank correlation. Intersections between lines and columns indicate the Spearman’s rank correlation coefficient (ρ) between the two parameters. Spearman ρ value is visualized by blue or red intensities. **B**: Correlation between relative levels of 2-AA-LPC and 25-hydroxycholesterol levels in carotid plaques (R and P values) were obtained by the Spearman rank test.

## Discussion

In this study, we examined the differences between T2DM and matched atherosclerotic patients in terms of plaque characteristics, lipid composition, biological parameters, systemic inflammation, and oxidative stress markers. Even within our specific cohort of elderly, poly-medicated patients with advanced atherosclerosis, we identified a characteristic diabetic signature. This included hyperglycemia, dyslipidemia (elevated TG and reduced HDL), oxidative stress (increased plasma 7α– and 7β-hydroxycholesterol), and heightened inflammation markers (hs-CRP and IL-1β) in the T2DM group. While no differences in plaque vulnerability or echogenicity between the groups was observed, we did find a significant enrichment of 2-AA-LPC in the plaques of diabetic patients, underlining the relevance of this molecule in the context of diabetic atheroma.

Our findings align with the existing literature that underscore the positive association of inflammation markers with the risk of diabetes.^25–27^ The association of hs-CRP and IL-1β with plasma triglyceride levels and BMI in both the general population and diabetic patients suggests that systemic inflammation plays a crucial role in the pathogenesis of diabetic atherosclerosis and is related to diabetic dyslipidemia. It is noteworthy that this heightened inflammation is apparent even in our study population, which comprises elderly patients with advanced atherosclerosis. While IL-1β was previously linked with the risk of diabetes, evidence for increased IL-1β levels is limited due to the typically low circulating levels of IL-1β. ^28–30^ It is therefore noteworthy that we could document here significantly higher IL-1β plasma concentrations in T2DM patients with advanced atherosclerosis.

We found mild differences in circulating oxysterol levels between diabetic and control patients, with only a moderate increase in 7α and 7β-hydroxycholesterol levels. Despite previous studies reporting increased plasma oxysterol levels in T2DM patients, this has not been consistently observed, especially when oxysterol levels were normalized by total cholesterol concentrations.^20, 31, 32^ Again, these differences may be attributed to the specificities of our study population, which warrants further investigation. Interestingly, we found no significant correlation between plasma oxysterol levels and their enrichment within the plaque, as observed previously.^22^ More generally, there was no relationship between blood biomarkers and the plaque molecular components. This suggests that the alterations in plaque composition are mainly driven by local metabolic and inflammatory processes and are not directly related to systemic markers.^33^

The relative increase in 2-AA-LPC in the plaques of diabetic patients observed in the present study is in line with our previous observation.^17^ Moreover, multivariate logistic regression analysis further supports the idea that 2-AA-LPC is independently associated with the risk of type 2 diabetes. 2-AA-LPC enrichment was not associated with systemic markers related to diabetes such as hsCRP, triglycerides or HDL cholesterol. To note, we previously observed a significant correlation of 2-AA-LPC with Hba1c ^17^ which is not retrieved here in a larger cohort of T2DM patients. Besides, 2-AA-LPC enrichment significantly correlated with inflammatory markers within the plaques, i.e. total LPCs, 27-hydroxycholesterol and 25-hydroxycholesterol, highlighting the potential role of inflammation in the development of diabetic atherosclerosis. Interestingly, the strong correlation between the levels of 25-hydroxycholesterol and the levels of 2-AA-LPC supports the hypothesis of related production pathways for the two molecules. A recent study has shown that 25-hydroxycholesterol accumulates in severe coronary artery lesions and demonstrated a specific role of 25-hydroxycholesterol production by macrophages to promote inflammation and vascular remodeling.^23^

Our study has some limitations. The cross-sectional design prevents us from establishing causal relationships between the observed changes and the development of diabetic atherosclerosis. Furthermore, the criteria for symptomatic lesions were based on the occurrence of CV events in the six months preceding surgery, and we lack post-surgery follow– up data on the occurrence of CV events. To confirm our findings and further explore the involved underlying mechanisms, longitudinal studies with larger sample sizes are necessary.

In conclusion, our study provides evidence for increased inflammation markers and enrichment of 2-AA– LPC in diabetic patients, strengthening the potential role of inflammation in the pathogenesis of diabetic atherosclerosis. While several inflammatory pathways could account for the increase production of 2-AA-LPC,^34–37^ further research is needed to better understand the local processes involved in the alterations of plaque composition in T2DM and to identify potential therapeutic targets for the prevention and treatment of diabetic atherosclerosis.

## Article information

All the data and details of analytical methods are available from the corresponding authors upon request.

## Data Availability

All data produced in the present study are available upon reasonable request to the authors

## Acknowledgments

The authors thank Hélène Choubley and Victoria Bergas from the lipidomic platform for excellent technical assistance.

## Sources of Funding

This work was supported by the University Hospital of Dijon Bourgogne and by grants from the Conseil Régional de Bourgogne Franche-Comté, by a French Government grant managed by the French National Research Agency under the program ‘Investissements d’Avenir’ with reference ANR-11-LABX-0021 (Lipstic Labex).

## Disclosures

The authors have no conflict of interest to disclose

## Notes

### Competing Interest Statement

The authors have declared no competing interest.

### Funding Statement

This work was supported by the University Hospital of Dijon Bourgogne and by grants from the Conseil Regional de Bourgogne Franche-Comte, by a French Government grant managed by the French National Research Agency under the program Investissements d Avenir with reference ANR-11-LABX-0021 (Lipstic Labex).

### Author Declarations

The Ethics Committee (Comite de Protection des Personnes Est, Dijon, France CPP Est III, CHRU Nancy, Number 2017-A00022-51) gave ethical approval of this work. This work was registered on ClinicalTrials.gov (clinical registration number: NCT03202823

